# Gender inequities regarding the HPV vaccination coverage in high-income countries: a systematic review and meta-analysis

**DOI:** 10.1101/2025.03.03.25323234

**Authors:** Sasidharanpillai Sabeena, Nagaraja Ravishankar

## Abstract

**Objective:** This systematic review aimed to systematically map the evidence and identify the knowledge gaps in the universal HPV vaccine coverage in high income countries.

**Methods:** An electronic search of the PubMed/MEDLINE, Scopus, Google Scholar, and official websites was carried out for articles reporting the HPV vaccination coverage in high income countries. The data was analysed systematically based on the updated Preferred Reporting Items for Systematic Reviews and Meta-analyses (PRISMA) checklist. The meta-analysis component was modified appropriately to synthesise the pooled prevalence of the HPV vaccine uptake among adolescents, men having sex with men and transgender persons with 95% confidence interval (CI). The random effects meta-analysis was performed in STATA version 13.0 (College Station, TX, USA) to evaluate and summarise the results. The forest plots were made using the midas package in STATA. The heterogeneity between qualified studies was reported using I^2^ index.

**Results:** Twenty-three articles regarding the HPV vaccination coverage were qualified for the quantitative synthesis including 27,80,307 adolescents, 11,909 men having sex with men and 207 transgender women. The pooled prevalence of the adolescent HPV vaccine uptake among adolescent girls was 67.75% (95% CI: 62.81%,72.69%) and adolescent boys was 58.32% (95% CI 48.88%, 67.76%). The pooled prevalence among men having sex with men (MSM) was 27.94% (95% CI 14.35, 41.53) and the pooled prevalence in transgender women was 32.59% (95% CI 8.35%. 56.83%)

**Conclusion:** Significant gender inequities exist in the HPV vaccination coverage and less number of gender minorities were vaccinated against HPV in high income countries. The major challenges were low awareness, lack of recommendations from clinicians, and public mistrust towards all vaccines.

**PROSPERO registration number:** CRD42025641689

**What is already known on this topic:**

➢ HPV vaccination programmes extending to adolescent boys and catch-up vaccination of men up to 26 years is more cost-effective than vaccinating only adolescent girls and women.
➢ The gender neutral HPV vaccination approach ensures gender equity as gender diverse populations such as TGW and MSM are at higher risk of persistent HPV infections.
➢ The HPV vaccination coverage is suboptimal among adolescents and young adults in high income countries, despite the well proven vaccine efficacy.

**WHAT THIS STUDY ADDS:**

➢ The rural health care providers were less likely to recommend adolescent HPV vaccination for cancer prevention.
➢ The HPV vaccine coverage of transgender women and men having sex with men (MSM) is minimal, despite being at higher risk of HPV infection.
➢ In contrast to TGW, men having sex with men in the USA showed better awareness of sexually transmitted diseases and were not hesitant to consult healthcare professionals if needed.
➢ The reduced HPV vaccine uptake among sexual minorities was mainly attributed to the low awareness among healthcare professionals, lack of recommendations by clinicians, and public mistrust of healthcare systems.

**HOW THIS STUDY MIGHT AFFECT RESEARCH, PRACTICE OR POLICY:**

➢ This systematic review presents the pooled prevalence of gender neutral HPV vaccination uptake that may assist policy makers, and stakeholders in making shared decisions.
➢ The barriers to HPV vaccination uptake should be managed by targeted public health and clinical practice interventions.
➢ Further research among gender minorities should be planned to analyse their perceptions and to bridge the knowledge gaps.

## Introduction

The Human Papillomavirus (HPV) infection is a sexually transmitted viral infection affecting all genders. World Health Organization (WHO) has recommended the HPV vaccination coverage of 90% among adolescent girls before the age of 15 years, screening of 70% of women by 35 years, and management of cervical pre-invasive lesions and cervical cancers in 90% of women for global elimination of cervical cancer [1]. However, a persistent rise in HPV-associated oropharyngeal squamous cell cancers (OPSCC) among men has been reported in high-income countries over the past few decades [2].

World Health Organization has listed the HPV vaccine as an essential medicine [3]. Gender neutral or universal HPV vaccination approach promotes vaccine acceptance, minimizes vaccine misinformation, and reduces vaccine-related stigma. The HPV vaccination should also be extended to gender diverse individuals to reduce the global burden of HPV-associated cancers. Even though many countries have implemented the HPV vaccination programme for women, gender-neutral HPV vaccine coverage is negligible [4]. HPV vaccination of adolescent boys and catch-up vaccination of men up to 26 years is more cost-effective than vaccinating only adolescent girls and women [5,6].

The countries implementing gender-neutral HPV vaccination are high-income or upper middle income countries except Bhutan [7]. Currently, forty-seven countries such as Australia, Canada, France, Ireland, Hong Kong, Portugal, South Korea, Switzerland, the United Kingdom, and the United States of America have introduced the gender neutral HPV vaccination in their national immunization programmes. The HPV vaccination coverage is suboptimal and varies widely depending on the gender identity or sexual orientation in high income countries. The gender neutral HPV vaccination approach ensures gender equity as gender diverse populations such as TGW and MSM are at risk of persistent HPV infection. They often encounter distinct barriers to healthcare and preventive vaccination services. The updated data regarding the human papillomavirus (HPV) vaccination coverage among adolescents, men having sex with men (MSM), and transgender individuals are essential to monitor the HPV vaccination programmes and analyse the impact of universal HPV vaccination on HPV-related disease burden.

This systematic review aimed to analyse the studies, systematically map the evidence and identify the knowledge gaps pertaining to the HPV vaccine uptake among adolescents, adult transgender persons, and MSM up to the age of 26 years in high income countries.

### Description of condition

#### HPV vaccine coverage

As per the WHO definition, HPV vaccine coverage is defined as the percentage of the target population (9-14 year old girls) who have received the recommended doses of HPV vaccine in a given year [8]. The uptake of single dose HPV vaccine was considered complete coverage based on the revised HPV vaccination recommendation in 2023 [9]. The US Advisory Committee on Immunization Practices (ACIP) has recommended gender neutral HPV vaccination across all genders up to the age of 26 [10]. The vaccine effectiveness decreases with increasing age.

#### HPV vaccine uptake

The HPV vaccine uptake indicates at least one dose of the HPV vaccine in adolescents, transgender persons, and MSM up to 26 years.

#### Transgender person

Transgender person is an “umbrella term” with no accepted definition and describes an individual whose sexual identity differs from the sex assigned at birth [11].

## 2. Methods

The review was started after ruling out any registered or ongoing systematic reviews concerning gender inequities of the HPV vaccination coverage in PROSPERO database. The study protocol was registered in the PROSPERO database (CRD42025641689) and may be accessed through the link https://www.crd.york.ac.uk/prospero/display_record.php?RecordID=641689. An electronic search of the PubMed/MEDLINE, Scopus, Google Scholar, and official websites was carried out. Articles published in English between January 2010 and January 2025 regarding the gender disparity in HPV vaccination coverage were screened. Relevant articles were identified electronically using specific search terms and related articles in peer-reviewed journals were manually searched. The references of retrieved articles were reviewed and the last date of the search was noted **(15^th^ February 2024)**.

Observational studies reporting the HPV vaccination coverage in adolescents and gender minorities were screened. The study population included vaccinated adolescents, MSM, and transgender individuals aged 18-26 years from a high income country. The studies reporting HPV vaccination coverage only among girls or boys from a specific region were not included. Cost-effectiveness studies, modelling studies, studies carried out among intellectually disabled adolescents, and migrant adolescents were excluded. Studies carried out among adults above 26 years were also not included.

### Data extraction

PubMed/MEDLINE, Scopus, and Google Scholar were searched for the articles published between January 2010 and 2015. The relevant articles in English were screened using search terms such as “adolescents” AND “Human Papillomavirus vaccines” OR “HPV vaccines” AND transgender” AND “MSM” OR “men having sex with men” AND “high income countries”.

A three-stage selection process was carried out for the final inclusion. One reviewer assessed the article titles and appropriate studies were moved to the second stage after eliminating irrelevant topics. In the second stage, the abstracts of the studies were independently assessed. After reviewing the abstracts, the full texts of the studies were retrieved which were scrutinised by two reviewers independently. The corresponding authors were contacted electronically if further interpretation was required. A manual library search for articles in peer-reviewed journals was carried out and references of retrieved articles were also reviewed to increase the search sensitivity. The study selection process was demonstrated in the PRISMA chart **(Figure 1)**. A validated proforma including the first author, year of publication, region, study design, study population, sample size, gender, study period, HPV vaccination coverage, and quality of the studies was prepared.

**Figure 1:**
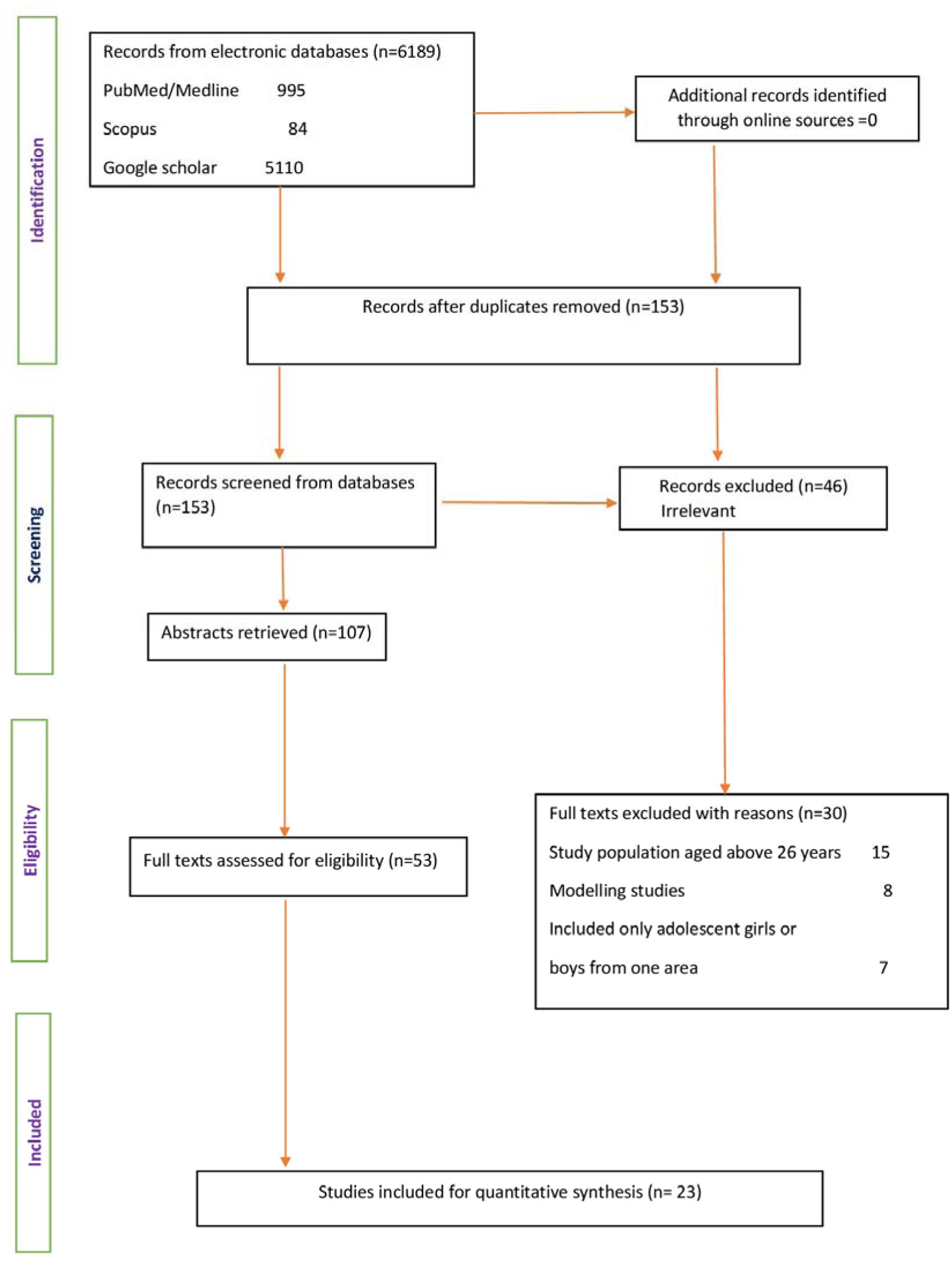
The PRISMA 2020 flow diagram for systematic reviews [41]. The flow diagram illustrates the number of articles screened, abstracts retrieved and full texts included/ excluded for the systematic review and meta-analysis. PRISMA: Preferred Reporting Items for Systematic Reviews and Meta-analyses.

### Risk of bias (quality) assessment

To assess the risk of bias in individual studies (quality assessment) identified after the screening, the National Institutes of Health (NIH) checklist for observational, cohort, and cross-sectional studies was used [12]. The studies with a minimum score of eight or above, seven, and five or below “Yes responses” were considered good, fair, and poor quality respectively.

For cross-sectional studies, question numbers 1, 2, 3, 4, 5, and 11 were relevant. The responses to the remaining eight questions (6-10, 12, 13, 14) were marked as not applicable (NA). The studies with six “Yes” responses were considered good and those with four /five were qualified as fair. The studies with less than four “Yes responses” were qualified as of poor quality. The quality of the studies was assessed independently by two reviewers.

### Meta-analysis

Meta-analysis was performed in STATA version 13.0 (College Station, TX, USA). The forest plots were made using the midas package in STATA. The pooled HPV vaccine uptake with a 95% confidence interval was reported among adolescent girls, adolescent boys, transgender women, and MSM up to the age of 26 years. As **s**ubstantial methodological and statistical heterogeneity was anticipated, the random effects model was applied. The forest plots depict the pooled prevalence of HPV vaccination uptake with 95% CI and Chi-square statistic (Q statistic). The heterogeneity between qualified studies was reported using I^2^ index. The values ranging between 25%-49%, 50-74%, and 75-100% imply low, moderate, and high heterogeneity respectively [13].

### Assessment of Publication Bias

Egger’s test was used to report the publication bias. Weighted linear regression with standardised effect estimate was the dependent variable and precision was the independent variable. In the present study, the log^e^ prevalence rate of HPV vaccine uptake among adolescents, transgender women, MSM, and precision were considered the effect estimate. The precision was reported as 1/standard error of log^e^ prevalence rate. Weights were allotted using the inverse variance approach (1/variance of the effect estimate). The statistically significant bias coefficient implies publication bias (*P <* 0.05).

## 3. Results

### 3.1 Identification and selection of sources of evidence

Using specific search terms, 153 articles from high income countries were screened from electronic databases and fifty-three articles were systematically reviewed (**Figure 1**). Twenty-three articles regarding the HPV vaccination coverage were qualified for the meta-analysis (quantitative synthesis) including eleven studies on gender neutral adolescent HPV vaccination [14–23] and six studies each among transgender women [24–29] and men having sex with men [26,27,30–33].

### 3.2 Characteristics of sources of evidence

Out of the eleven qualified studies on adolescent HPV vaccination uptake, five studies from the USA, two from Australia, and one each from Canada, Austria, France, and Germany were analysed **(Table 1)**. The study population included 27,80,307 adolescents including 19,75,425 girls and 8,04, 882 boys. All these studies were cross sectional studies except three cohort studies from Germany, Canada, and France [20,21,23]. The HPV vaccination uptake varied between 46%-97.4% among adolescent girls. Meanwhile, the uptake of the HPV vaccine varied between 27.9%-95.3% among adolescent boys. All the six qualified studies reporting the HPV vaccination coverage among 207 transgender women (TGW) were from the USA **(Supplementary Table 1)**. The study sample of TGW reporting to health clinics ranged between six to sixty-five. The HPV vaccination coverage among the study population varied between 4.3%-88%. The HPV vaccination data among 11,909 MSM were retrieved from six US studies **(Supplementary Table 2)**. Three studies enrolled young men from twenty metropolitan statistical areas as part of National HIV Behavioural Surveillance (NHBS).[30,31,33] Meanwhile, two qualified studies were from the multicentre Vaccine Impact in Men (VIM) study [26,27]. Except for one study by Thomas et al, all the qualified studies included adult MSM [32]. Thomas et al included data from young boys aged 13 to 18 years and adults up to the age of 26 years attending a sexually transmitted disease clinic and teen health centre. The number of MSM attending the health clinics ranged between four hundred to three thousand one hundred and fifty-five. The HPV vaccination coverage varied between 4.9% to 47.6%. Quality of all the studies included for the meta-analysis was good.

**Table 1:** Table depicts the characteristics of qualified studies on gender neutral HPV vaccination uptake in adolescents.

| No | Author (year) | Country | Study population | Age group | Study design | Study period | Sample (N) | Vaccination uptake Girls N (%) | Vaccination uptake Boys N(%) | Limitations | Quality |
| --- | --- | --- | --- | --- | --- | --- | --- | --- | --- | --- | --- |
| 1 | Singer et al. (2025) | Bremen, Germany | School students | 13-14 | Cohort study | 2022-2023 | **2694**<br>**Girls-1418**<br>**Boys-1276** | **647 (46)** | **361(28)** | COVID-19 and Ukraine war contributed to lower response rate. Small cohort size. Data from limited period was considered. | Good |
| 2 | McIndoe et al. (2024) | Australia | School students | 11-12 | Cross sectional | 2019 | **9379**<br>**4572 girls**<br>**4807 Boys** | **4023 (88)** | **4134 (86)** | Findings may not be generalizable, as the study was conducted in south eastern Sydney. Catch-up and general practitioner administered HPV vaccination coverage were not included. | Good |
| 3 | Crystal Du et al. (2024) | Alberta, Canada | non-immigrant adolescents | 12-18 | Cohort | 2014-2018 | **208, 2 56**<br>**101,725 girls**<br>**106,531 boys** | **64,606 (63.51)** | **42,357 (39.76)** | Coverage data was limited to Western Canadian Province and may not be generalizable. | Good |
| 4 | de Pouvoirville et al.(2024) | France | adolescents | 11-19 | Cohort Claims study | 2017-2022 | **2,366,067**<br>**Girls-1,773,900**<br>**boys 592,167** | **1,193,834 (67.3)** | **369,512 (62.4)** | Number of vaccines dispensed was taken as actual vaccination data resulting in overestimation. Vaccinated during free medical visits was not recorded in the national database leading to underestimation. | Good |
| 5 | Boakye et al. (2022) | Illinois USA | adolescents | 11-17 | Cross sectional | 2015-2020 | **9351**<br>**girls 4649**<br>**boys 4702** | **2241(48.2)** | **2076 (44.1)** | Data from one medical centre was analysed. Hence may not be representative of the national situation. | Good |
| 6 | Pingali et al. (2021) | USA | adolescents | 13-17 | Cross sectional study NIS-Teen survey | 2020 | **20,163**<br>**girls 9,576**<br>**boys 10,587** | **7383 (77.1)** | **7739 (73.1)** | Low response rate may increase the bias. As per the survey error assessment, NIS Teen survey might underestimate true coverage. Households without telephone could not be included. | Good |
| 7 | Elam Evans et al. (2020) | USA | adolescents | 13-17 | cross sectional study<br>NIS-Teen survey | 2019 | **18,788**<br>**girls 8916**<br>**boys 9872** | **6526**<br>**(73.2)** | **6890**<br>**(69.8)** | Lower response rate due to COVID-19 pandemic.<br>Increase in bias rates.<br>Phone-less households could not be included. | Good |
| 8 | Swiecki-Sikora et al. (2019) | USA | adolescents | 13-17 | cross sectional study<br>NIS-Teen survey | 2012-2013 | **37,114**<br>**Girls-17,596**<br>**Boys-19,518** | **9783**<br>**(55.6)** | **5,445 (27.9)** | Low response rate and minimal data were provider verified.<br>Inclusion of provider verified records would have caused underestimation. | Good |
| 9 | Walker et al. (2018) | USA | adolescents | 13-17 | cross sectional study<br>NIS-Teen Survey | 2017 | **103,703**<br>**girls 50,711**<br>**boys 52,992** | **34,788**<br>**(68.6)** | **33,172**<br>**(62.6)** | Biases in estimates due to low response rates. | Good |
| 10 | Brotherton et al (2017) | Australia | adolescents | 12 years | Cross sectional study pf<br>NHPVR | 2015 | **4,343**<br>**Girls- 2108**<br>**Boys- 2235** | **2054**<br>**(97.4)** | **2129**<br>**(95.3)** | HPV vaccination by general practitioners might not be notified resulting in underestimation. | Good |
| 11 | Borena et al. (2014) | Austria | adolescents | 9-10 | cross sectional | 2015 | **449**<br>**girls 254**<br>**boys 195** | **150 (59)** | **101 (51.8)** | Data from limited number of adolescents from western region<br>adolescents from rural areas not included. | Good |

### 3.3 Meta-analysis (Quantitative synthesis)

The pooled prevalence of the HPV vaccine uptake among 19,75,415 adolescent girls from eleven qualified studies was 67.75% (95% CI: 62.81%,72.69%). As shown in the **Figure 2** the I^2^ value was 99.92%. The pooled prevalence of the HPV vaccine uptake among 8,04,882 adolescent boys was 58.32% (95% CI 48.88%, 67.76%) with an I^2^ value of 99.98% (**Figure 3**). The pooled prevalence of the HPV vaccine coverage among 207 transgender women was 32.59% (95% CI 8.35%. 56.83%) with an I^2^ value of 96.16% **(Figure 4).** The pooled prevalence of the HPV vaccine uptake among 11,909 men having sex with men (MSM) was 27.94% (95% CI 14.35, 41.53) and I^2^ value was 99.72% **(Figure 5)**.

**Figure 2:**
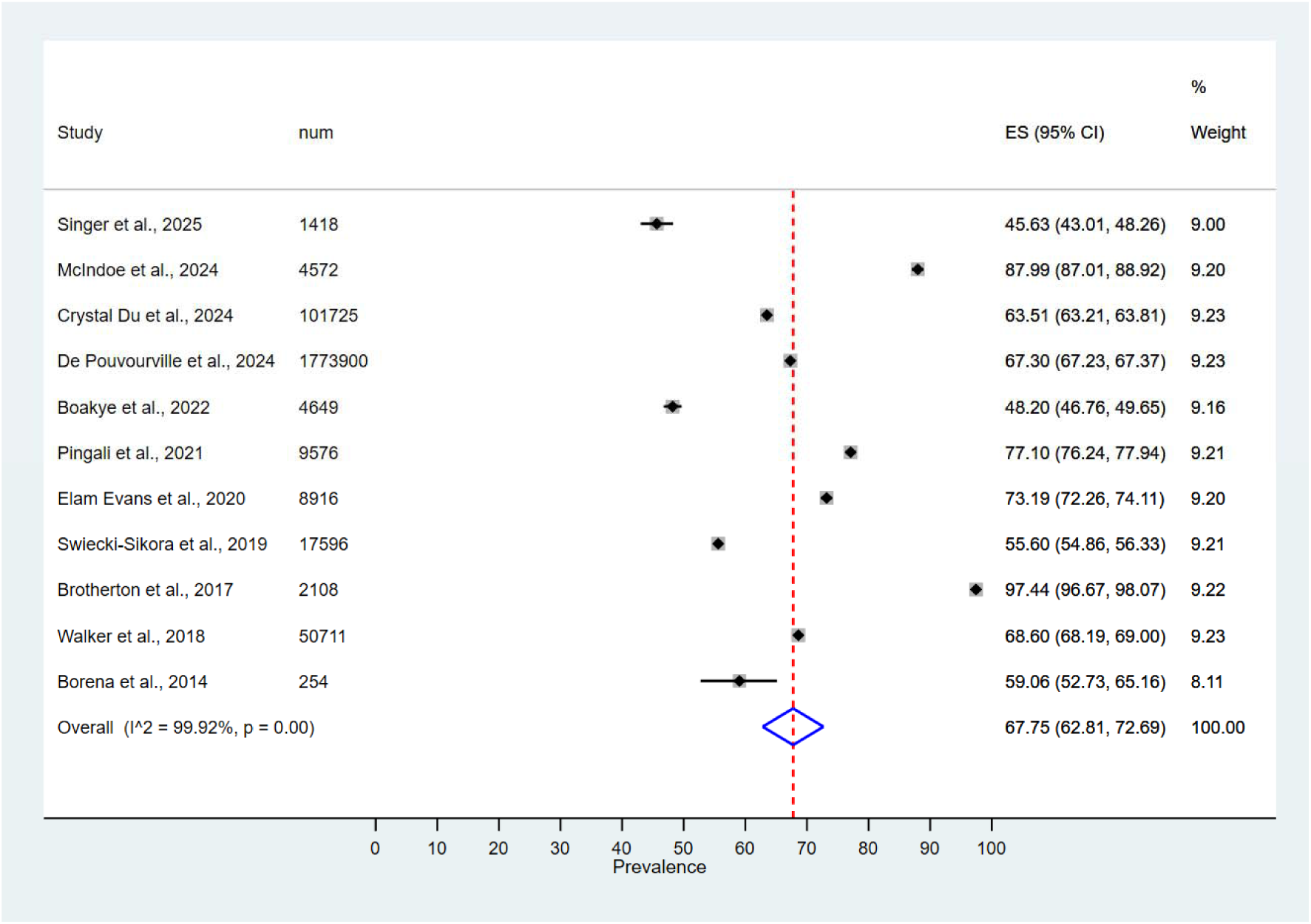
Forest plot of the summary effect size (pooled prevalence of HPV vaccine uptake among adolescent girls) using random effects model. Squares imply the effect size of individual studies and extended lines represent 95% confidence interval. The size of the squares indicate the weight of the studies based on sample size using a random effects analysis. The diamond data indicates the pooled prevalence. Test of heterogeneity = 99.92%, *P*-value =0.00. CI confidence interval.

**Figure 3:**
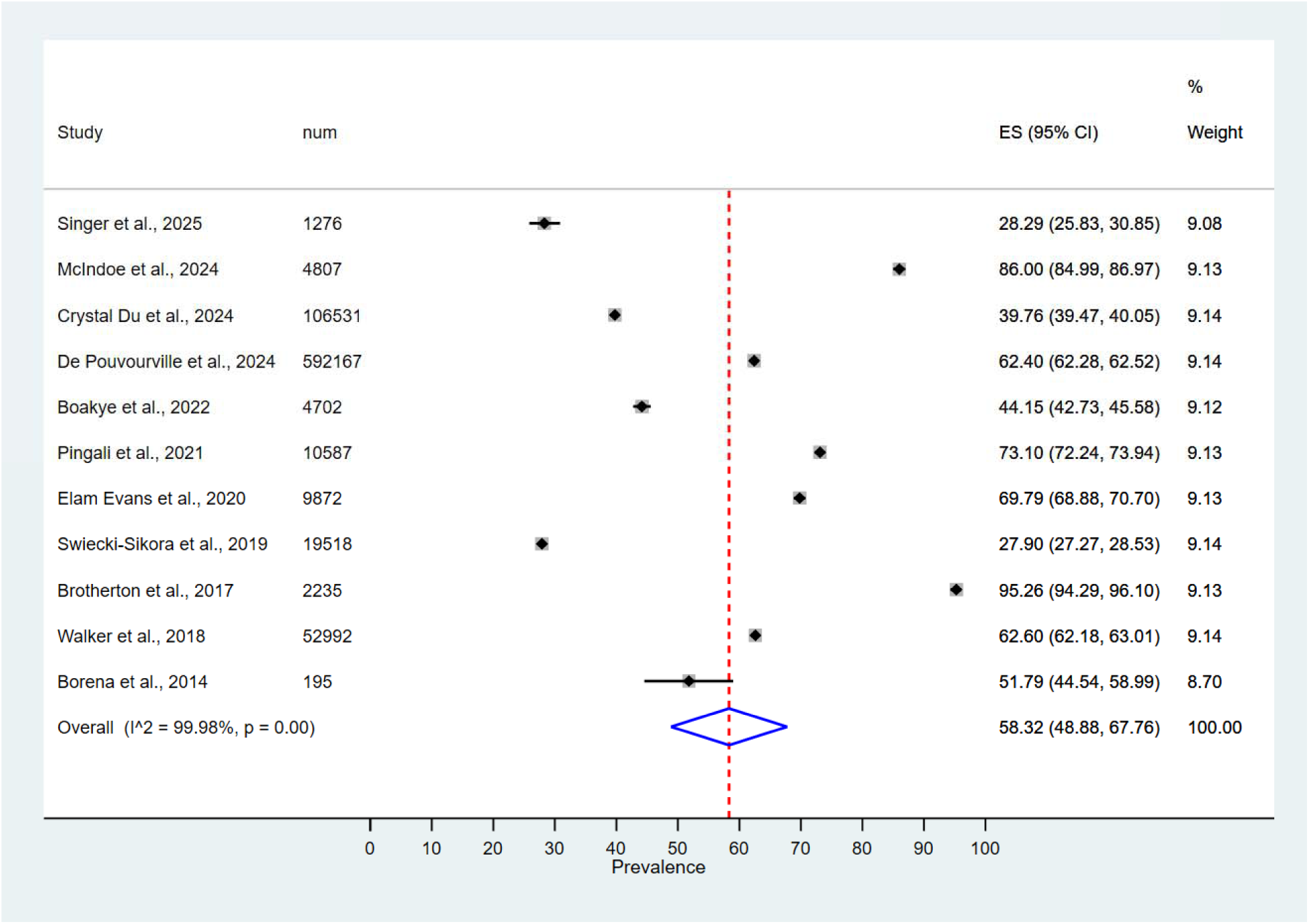
Forest plot of the summary effect size (pooled prevalence of HPV vaccine uptake among adolescent boys) using random effects model. Squares imply the effect size of individual studies and extended lines represent 95% confidence interval. Size of the squares indicate the weight of the studies based on sample size using a random effects analysis. The diamond data indicates the pooled prevalence. Test of heterogeneity =99.98%, *P*-value = 0.00. CI confidence interval.

**Figure 4:**
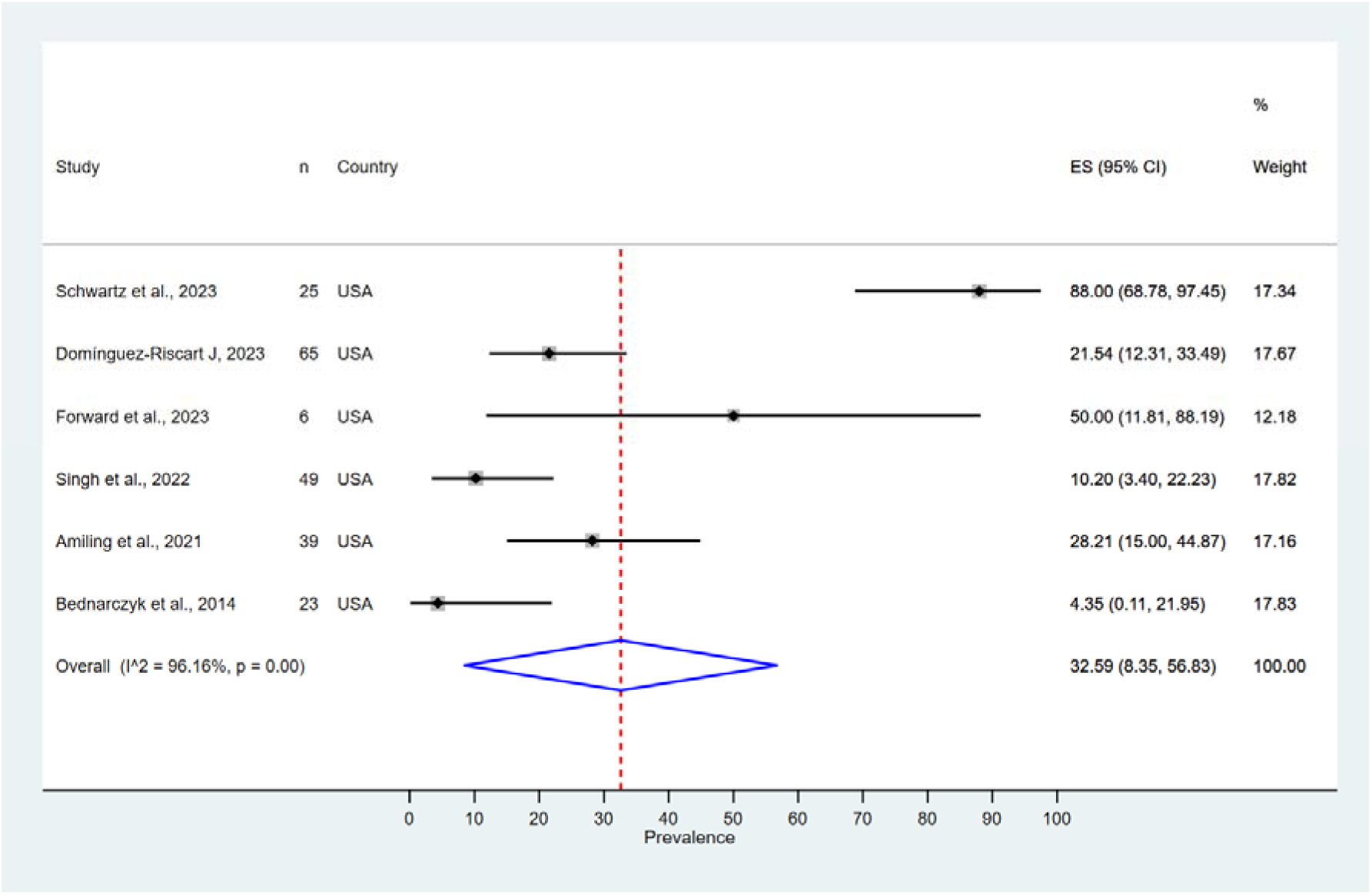
Forest plot of the summary effect size (pooled prevalence of HPV vaccine uptake among transgender women) using random effects model. Squares imply the effect size of individual studies and extended lines represent 95% confidence interval. Size of the squares indicate the weight of the studies based on sample size using a random effects analysis. The diamond data indicates the pooled prevalence. Test of heterogeneity =96.16 %, *P*-value =0.00. CI confidence interval.

**Figure 5:**
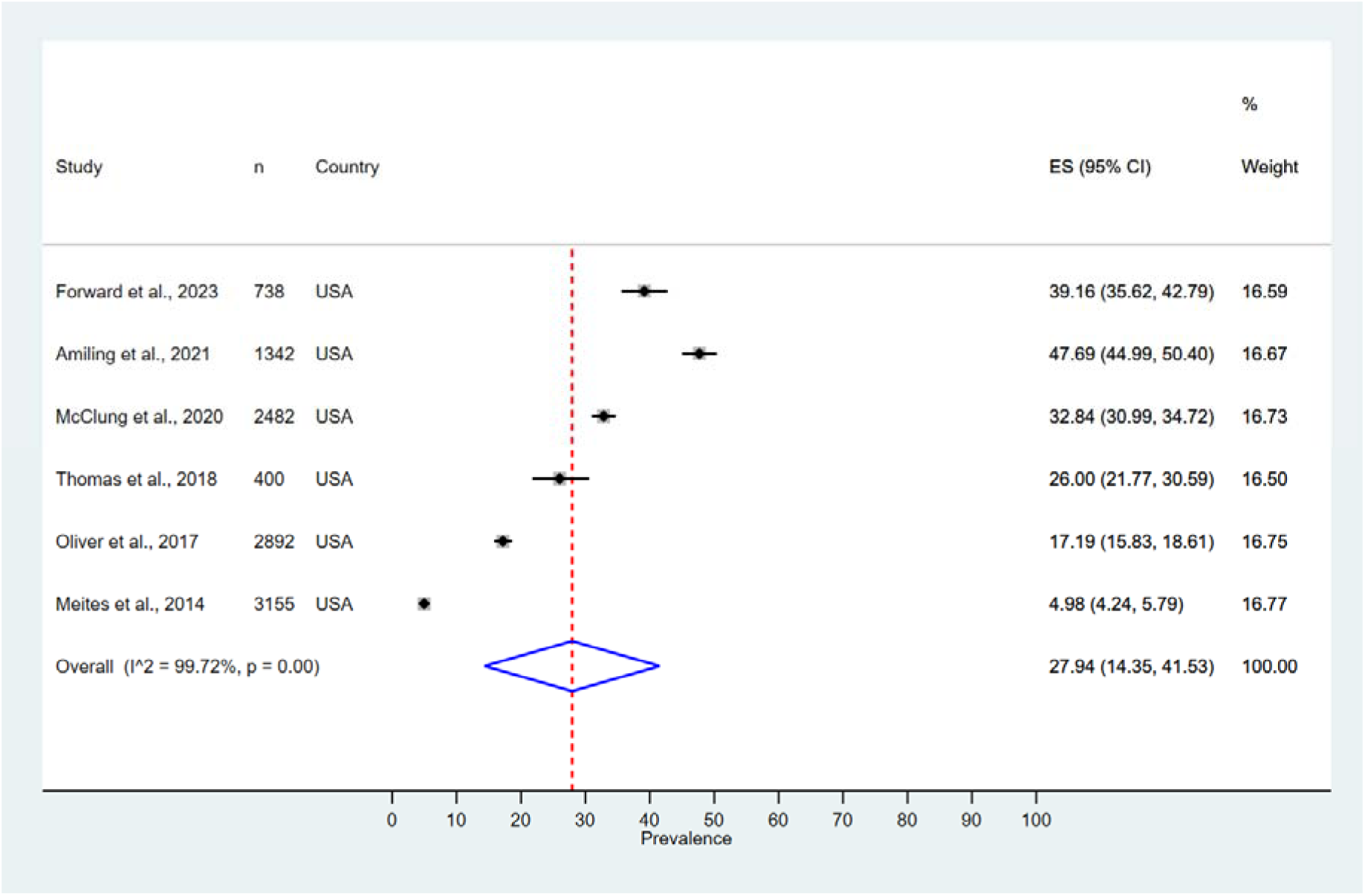
Forest plot of the summary effect size (pooled prevalence of HPV vaccine uptake among men having sex with men) using random effects model. Squares imply the effect size of individual studies and extended lines represent 95% confidence interval. Size of the squares indicate the weight of the studies based on sample size using a random effects analysis. The diamond data indicates the pooled prevalence. Test of heterogeneity =99.72 %, *P*-value =0.00. CI confidence interval

#### Publication bias

The *p*-value for the bias coefficient is above 0.05, which is statistically non-significant. There is no evidence of publication bias (**Supplementary Table 3**).

## Discussion

There is evidence of gender disparity in the HPV vaccination coverage in high income countries. The present review systematically analysed the HPV vaccination coverage among adolescents, TGW, and MSM. Persistently high HPV vaccine coverage was observed among adolescent girls in all the qualified studies. Meanwhile, the HPV vaccination coverage was suboptimal among adolescent boys, MSM and TGW. Transgender women and MSM were enrolled from sexually transmitted disease (STD) centres, gender diversity centres, or HIV surveillance units. Hence, the study findings may not be applicable to all the gender diverse persons in the USA. The number of TGW seeking the medical care was minimal with low intention to take the HPV vaccine. The prevalence of HPV infection was higher among TGW than among men having sex with men [25]. The reduced HPV vaccine uptake among sexual minorities was attributed to the low awareness among healthcare professionals, lack of recommendations by clinicians, and public mistrust of healthcare systems [34]. Meanwhile, MSM in the USA were more aware of sexually transmitted diseases in comparison to TGW and were not hesitant to consult healthcare professionals if needed [30,31]. Singh et al reported a higher HPV prevalence rate among TGW than MSM in the USA [25].

Generally, the rural health care providers were less likely to recommend adolescent HPV vaccination for cancer prevention [35]. Transgender women are not often included in epidemiological studies as they encounter many barriers and challenges in accessing appropriate healthcare. There is sparse information regarding the HPV vaccination uptake among gender minorities. Transgender individuals in the USA encounter higher morbidity and mortality than cisgender individuals [36]. As per the Swedish population based cohort study, sex reassigned transgender individuals demonstrate a higher mortality rate than the general population [37]. A higher incidence of sexually transmitted infections such as gonococcal, chlamydial, and Human Immunodeficiency Virus (HIV) infections were reported among transgender men and women [38]. The incidence of HPV-related cervical and anogenital cancers was more among gender diverse persons [39].

Most of the published studies were carried out among adolescents before the COVID-19 pandemic with adequate response rates. All the included studies were qualified as good. Except for one study conducted at a medical centre, all the retrieved US studies on adolescent HPV vaccination reported provider verified data from the National Teen Survey (NIS-Teen). The studies from high income countries in the Europe, Australia, and the USA were included. Across Europe, higher vaccine hesitancy was reported in France and Germany due to the lack of trust in healthcare system and low vaccine confidence [40]. This meta-analysis has included one study each from France and Germany to pool the adolescent HPV vaccination data.

## Limitations

Few studies were conducted during the pandemic with a low response rate due to COVID-19 related public health preventive measures. Both the COVID-19 pandemic and Ukraine war affected the response rate in a study from Germany [23]. The adolescent HPV vaccines administered by the general practitioners were not notified in many high income countries resulting in underestimation. Data among rural adolescents were lacking in some studies. Most of the US publications regarding adolescent vaccinations were based on the NIS Teen survey which collected information through telephone interviews. The households without telephones were not included in the survey underestimating the actual HPV vaccination coverage. As the major proportion of reviewed studies were cross-sectional studies, confounding factors such as provider recommendation could not be adjusted and causal inference could not be reported.

All the qualified studies regarding the HPV vaccination coverage among TGW and MSM were from the USA. The sample size was small in studies enrolling transgender women, resulting in wider confidence interval while pooling the data. Hence the findings may not be generalisable to the entire US. Few studies restricted the inclusion to individuals having English language expertise. Recruitment of TGW from rural areas through social media platforms would have caused selection bias. Self-reporting of HPV vaccination status by adult men having sex with men and TGW would have generated recall bias and social desirability bias. The studies about other gender diverse groups such as transgender men, and lesbians were not qualified.

## Conclusion

Significant gender inequities exist in the HPV vaccination initiation. The barriers to HPV vaccination uptake should be managed by targeted public health and clinical practice interventions. The major barriers were low awareness among healthcare providers and parents, lack of recommendations from trusted clinicians, and public mistrust towards all vaccines.

## Recommendations

1. Policymakers should promote gender inclusive HPV vaccination services to all sexually naïve adolescents and young adults, irrespective of their gender identity.
2. Partnerships between health clinics and gender diverse units or organisations should be established to assist gender minorities by disseminating knowledge.
3. Personalised care aiming for tailored interventions to meet the patient preferences in an appropriate social context should be adopted.
4. Patient-physician shared decision making should be promoted in the HPV vaccination programmes.

## Data Availability

All data produced in the present study are available upon reasonable request to the authors.

https://www.crd.york.ac.uk/prospero/display_record.php?RecordID=641689

## Acknowledgement

The pre-print was published in MedRxiv (MEDRXIV/2025/323234) on 7^th^ March 2025.

https://www.medrxiv.org/content/10.1101/2025.03.03.25323234v1

https://www.medrxiv.org/cgi/content/short/2025.03.03.25323234v2).

## Ethics statement

The author states that this manuscript does not involve any misconduct such as plagiarism, forgery, tampering, improper signature, multiple submission, repeated publication, split publication, etc. The systematic review and metalJanalysis used published data from peer-reviewed articles in indexed journals. As such, ethical approval to conduct the analysis was not sought.

## Data availability statement

The data that support the findings of this study are available from the corresponding author upon reasonable request (Dr Sasidharanpillai Sabeena).

All authors attest they meet the ICMJE criteria for authorship

## Author contributions

Conceptualization, Data curation: S Sabeena

Formal analysis: S Sabeena, N Ravishankar

Methodology: S Sabeena

Validation: S Sabeena & N Ravishankar

Original draft; S Sabeena

Writing/Editing – S Sabeena & N Ravishankar

---

**Supplementary Table 1:**
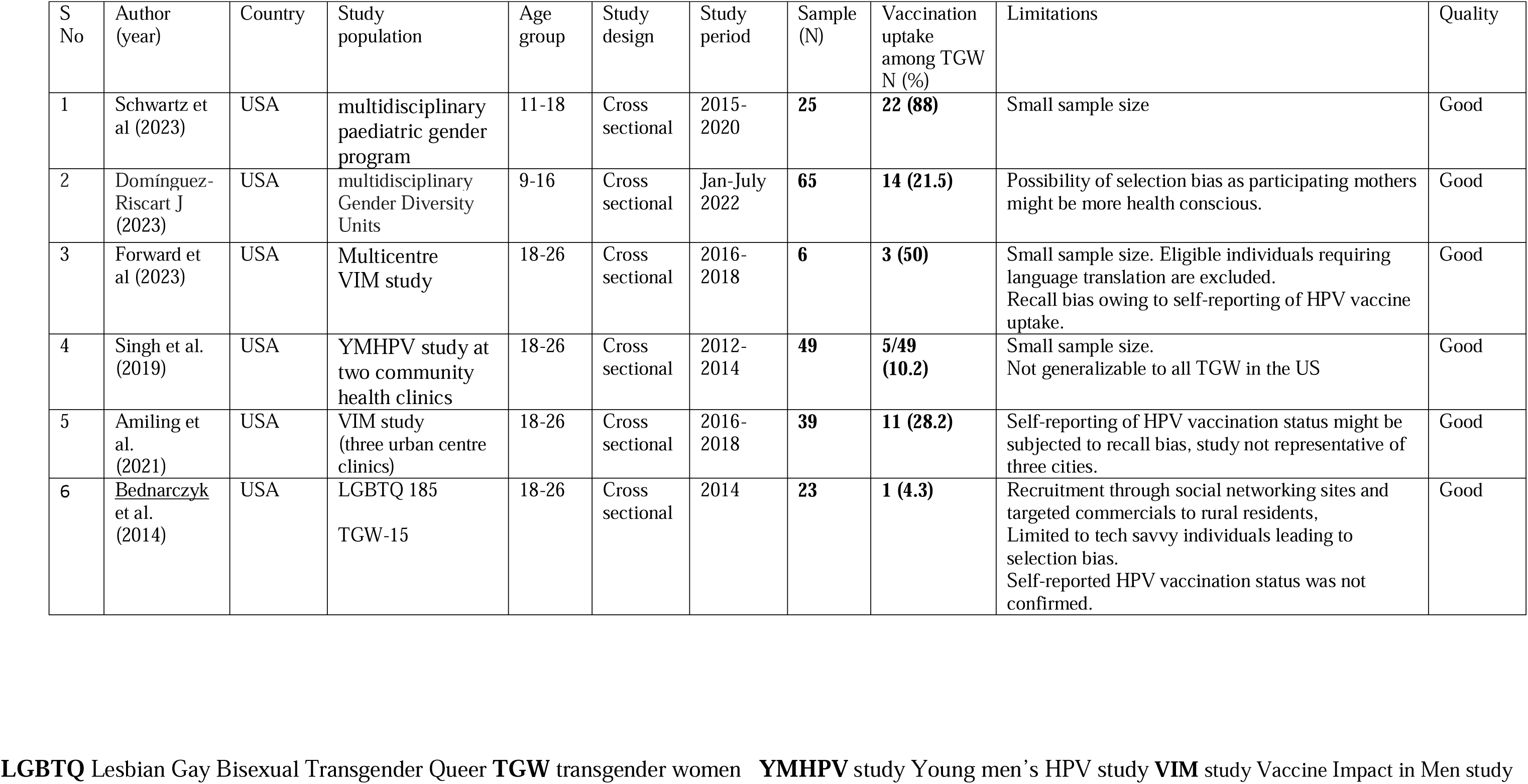
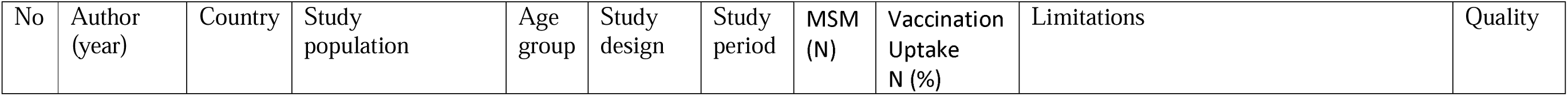
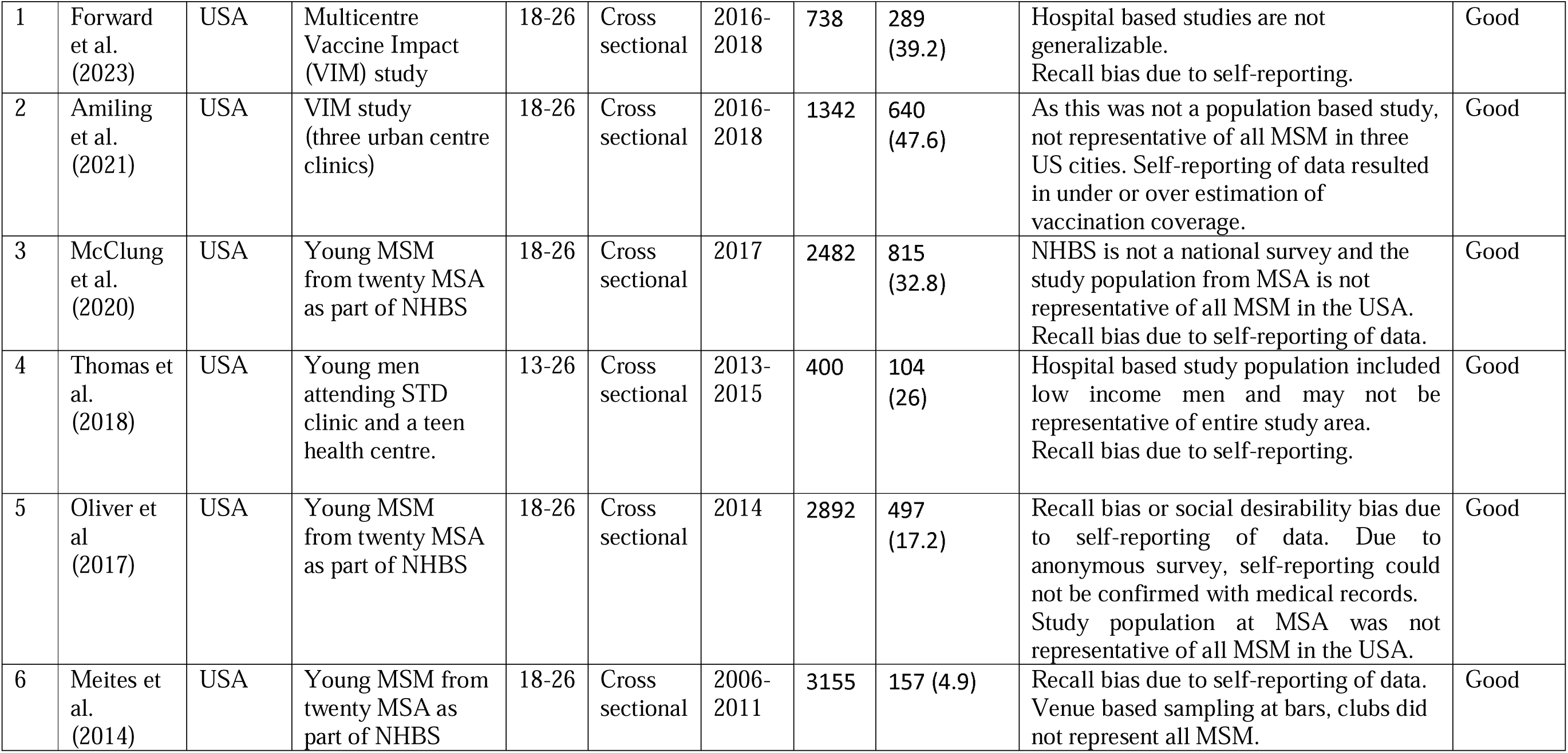
Characteristics of studies among Transgender women (TGW)

**Supplementary Table 3:** Table illustrates the publication bias.

**Girls**
| Coefficient | Estimate (95%CI) | P-value |
| --- | --- | --- |
| Slope | 0.6699 (0.6368, 0.7031) | <0.001 |
| Bias | 7.9543 (-22.1099, 38.0186) | 0.564 |

P-value for bias is not significant. This implies that publication bias is not present

Boys
| Coefficient | Estimate (95%CI) | P-value |
| --- | --- | --- |
| Slope | 0.5996 (0.4899, 0.7092) | <0.001 |
| Bias | -4.5722 (-66.4847, 57.3402) | 0.871 |

P-value for bias is not significant. This implies that publication bias is not present

TGW
| Coefficient | Estimate (95%CI) | P-value |
| --- | --- | --- |
| Slope | -0.0991 (-0.0988, 0.7899) | 0.772 |
| Bias | 6.2439 (-9.6846, 22.1725) | 0.338 |

P-value for bias is not significant. This implies that publication bias is not present

MSM
| Coefficient | Estimate (95%CI) | P-value |
| --- | --- | --- |
| Slope | -0.0371 (-0.2068, 0.1325) | 0.576 |
| Bias | 27.6900 (-4.8353, 50.5447) | 0.078 |

P-value for bias is not significant. This implies that publication bias is not present

